# Creation of novel pediatric academic curriculum and its evaluation using mixed methods

**DOI:** 10.1101/2022.08.07.22277912

**Authors:** Martha Balicki, Darja Barr, Robert Renaud, Atul Sharma, Celia Rodd

## Abstract

**Introduction:** The Royal College of Physicians and Surgeons of Canada and the American Accreditation Council for Graduate Medical Education require resident skills in Evidence-Based Medicine and participation in research activities. Our first-year pediatric residents (PGY1s) were required to attend a novel, call-protected, 4-week Academic Skills and Knowledge (ASK) rotation to improve their skills as consumers of medical literature. Objectives of the study were to describe this curriculum and summarize its mixed-methods evaluation.

**Methods:** After 14 months of curriculum development, three annual cohorts of PGY1s wrote identical pre- and post-ASK quizzes (2017-19). In 2018 and 2019, we assessed knowledge retention with PGY1s re-writing the quiz after 6 months. Mean test scores were compared using paired t-tests. In 2017, pre- and post-ASK focus groups assessed resident feelings about the rotation.

**Results:** All eligible PGY1s (n=32) participated. Mean exam scores demonstrated increased knowledge (time0 mean±SD 52.6±11.0%; vs. time1 80.2±9.0%, p <0.001). Knowledge retention at 6 months was intermediate (time2 70.2±12.0%; time0 vs time2 p<0.001). In the pre-rotation focus group, residents looked forward to ASK; goals centered around growing from learner to expert. Post-ASK, residents were very satisfied. Resident participation in our annual Research Institute poster competition increased linearly from 0% in 2014 to 8% in 2020 (r=0.74, p=0.01).

**Discussion:** The ASK curriculum was successfully implemented, and increased knowledge persisted over time. Residents were satisfied with ASK and appreciated the structured curriculum building on core knowledge that they could immediately apply to their clinical work.

**Statements and Declarations:** All authors contributed to the study conception and design. Material preparation, data collection and analysis were performed by Martha Balicki, Darja Barr, Atul Sharma and Celia Rodd. The first draft of the manuscript was written by Martha Balicki and all authors commented on previous versions of the manuscript. All authors read and approved the final manuscript.

**Financial interests:** None of the authors have any relevant financial or non-financial interests to disclose. The authors did not receive support from any organization for the submitted work.

**Data availability:** The datasets generated during and/or analysed during the current study are available from the corresponding author on reasonable request.

## Introduction

Both the Royal College of Physicians and Surgeons of Canada and the Accreditation Council for Graduate Medical Education require resident skills in Evidence-Based Medicine (EBM) and participation in scholarly activities[1,2]. For both residents and faculty, teaching, assessment, and application of these skills has been challenging without sufficient support, explicit training, or dedicated time. Several approaches have been implemented[3–9] (see systematic reviews[10–12]).

To develop academic skills - including evaluation of scientific literature, multilevel learner teaching, quality improvement (QI), pediatric ethics, feedback, and research skills (e.g. study design, biostatistics) - a mandatory, full-time, call-protected, 4-week rotation was added in 2016 to the first 4 months of the pediatric residency (PGY1) at the University of Manitoba. All PGY1 trainees (n=10-12/year) were liberated from clinical duties and enrolled in our “Academic Skills & Knowledge” (ASK) curriculum. About 50 clinical and research staff acted as small-group preceptors, allowing residents to become aware of scholarly endeavours across the Department.

Sessions generally involved an introductory didactic component combined with hands-on application of the material, typically reinforced with homework. Attendance was mandatory and exercises needed to be satisfactorily completed to pass the rotation. Exercises were designed to mimic real world experiences encountered during residency and practice. See Table 1 for curriculum and exercises.

**Table 1:**
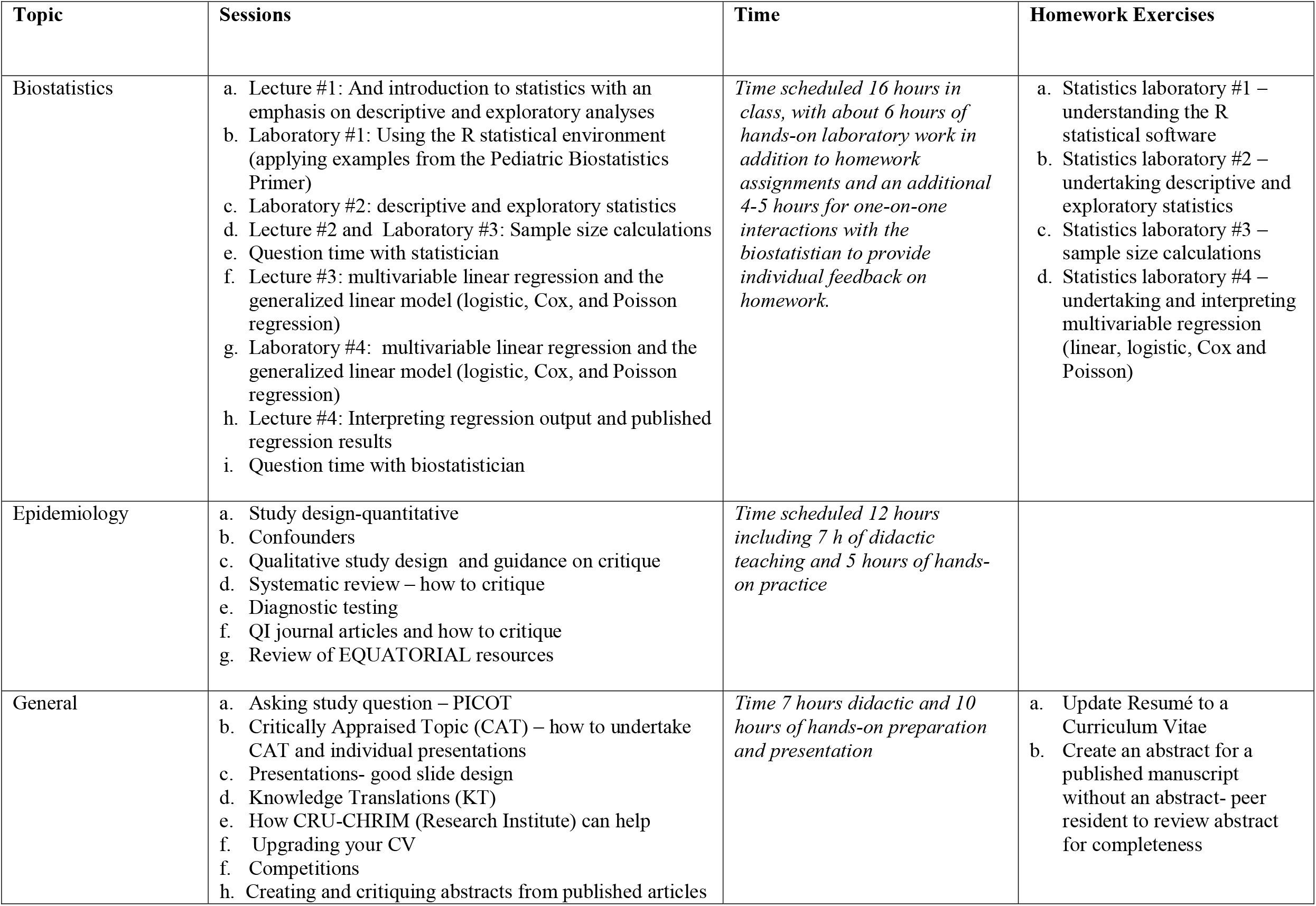
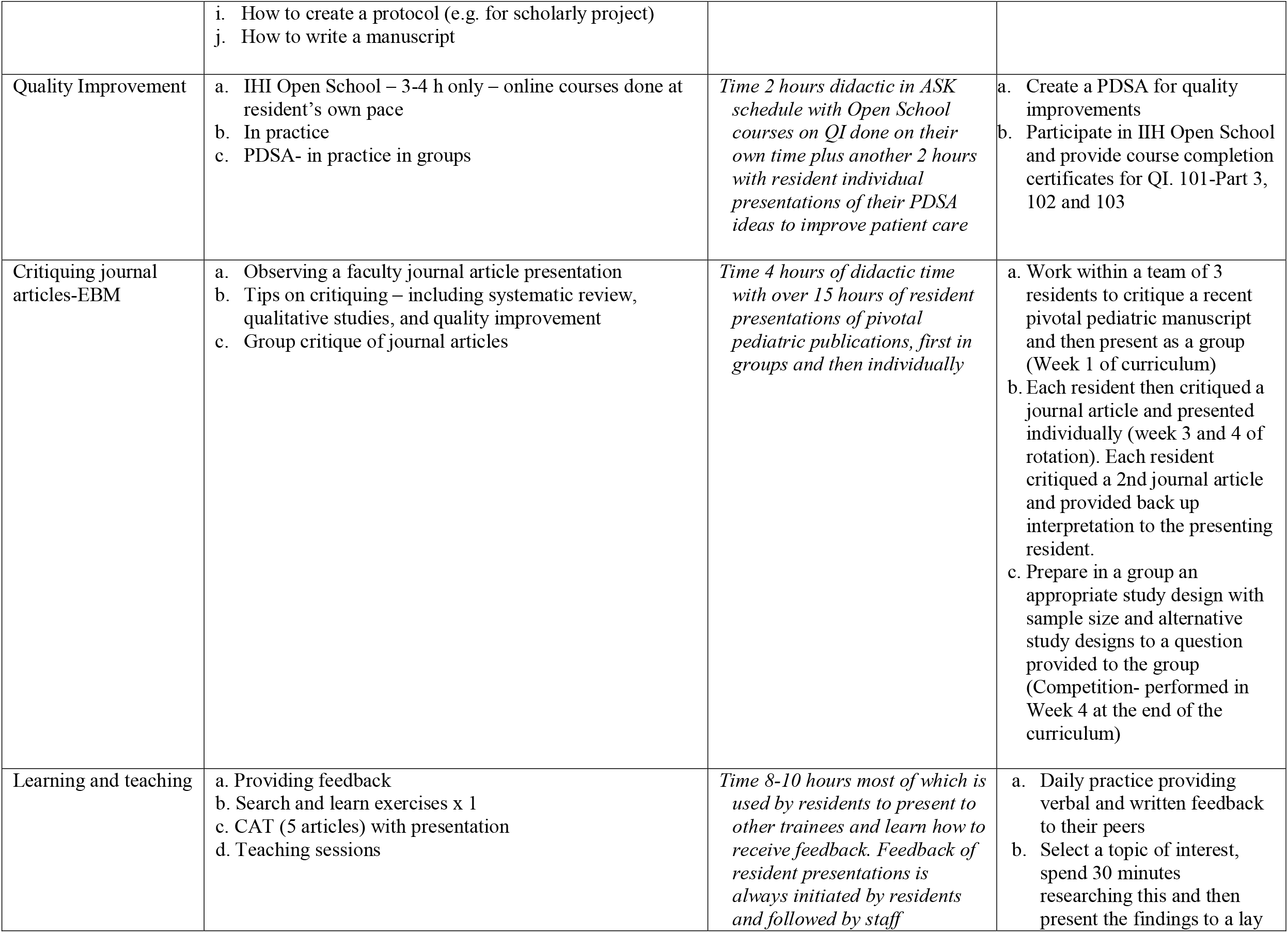
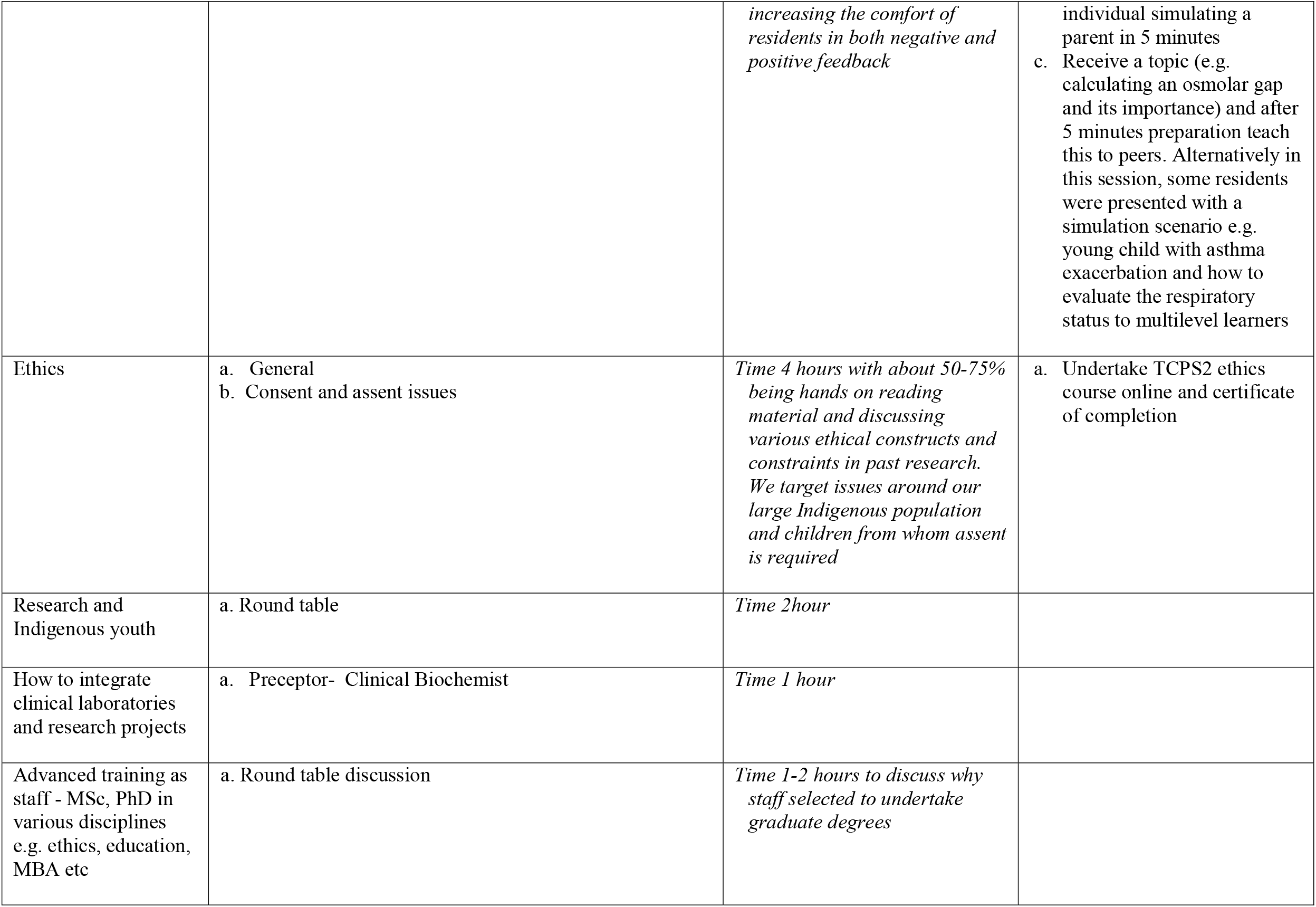
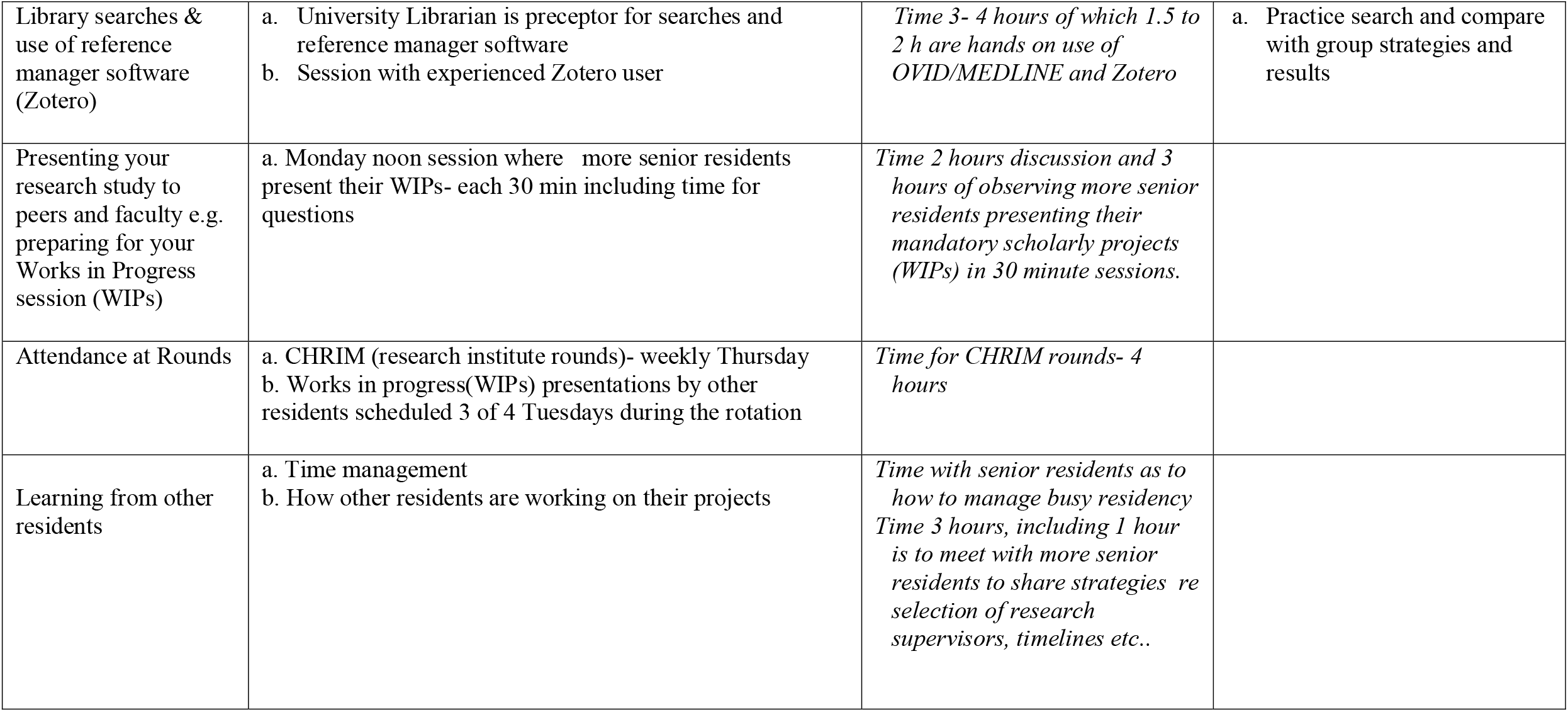
List of topics and homework exercises covered in ASK.

ASK expanded on an earlier lecture series spread over 4 years, with content covering study design and biostatistics to support their scholarly activities (e.g. journal clubs, research). Residents regularly missed sessions, and additional training was needed to support mandatory scholarly projects, which suffered from poor completion rates. Moreover, the Department wished to enhance residents’ skills as consumers of the pediatric literature early-on to augment clinical care.

Here we share our curriculum framework and resources, including our online Pediatric Biostatistics Primer and share the results of ongoing curriculum evaluation. We measured knowledge gains quantitatively with pre- and post-course quizzes and a knowledge retention exam 6 months post-course, which included a small control group of senior residents never exposed to ASK. We monitored resident scholarly productivity in terms of participation in our annual Children’s Hospital Research Institute research days. Lastly, we share insights from 2 focus groups assessing expectations and course feedback. We believe these resources and results will assist other programs implementing an academic curriculum.

## Materials and Methods

### Curriculum development

Our curriculum was structured using the 6 steps of Kern[13]

#### Step 1: Problem identification and general needs assessment

The senior author is a member of the Canadian Pediatric Resident Research group; bi-annual meetings are held with the Canadian Paediatric Society to share strategies to enhance scholarly and academic activities[14,15]. An extensive literature search was conducted, and contacts were made with other training programs to share curricula, which largely focussed on scholarly activities rather than the range of academic skills we wished to highlight.

#### Step 2: Targeted Needs Assessment

An ASK committee was formed within our Pediatric Department and University. This included experts in scholarly activities, biostatistics, QI, and education (under- and postgraduate), along with program directors and chief residents.

#### Step 3: Objectives and curricular goals

See Appendix A for objectives and goals in CanMEDS format. In July, 2021, Canada introduced Competency by Design (CBD) with Entrustable Professional Activities (EPAs) so these were not included in the original course design[16].

#### Step 4: Educational strategies

Our curriculum consisted of interactive seminars, independent reading activities, and internet-based courses (e.g. IHI Opens School, ethics training[17–19]). In 2017, 2 co-authors (AS and CR) created an online Biostatistical Primer teaching statistics through pediatric examples and hands-on laboratory session in the free R statistical environment. The primer, datasets and code are freely available from our <u>Research Institute</u> (https://www.chrim.ca/biostatistics/). Seminars and didactic sessions included case scenarios allowing for small group discussions. See Table 1 for more details.

#### Step 5: Implementation

The first course ran in fall 2016 for 4 weeks. Autumn was selected to allow the residents to complete 3-4 clinical rotations before immersion in ASK. Sessions were videotaped and were made available if needed for absence or review. Reading materials were provided as online pdfs for all presentations.

#### Step 6: Evaluation and Feedback

Written feedback was collected for each session and preceptor from all trainees; residents provided written and immediate verbal feedback on all peer presentations. Similarly, preceptors provided written feedback about residents. The ASK committee had access to anonymized resident feedback when they met to revise for subsequent years, with one first year resident added to the group annually. The 2016 curriculum was trimmed slightly in length (the course runs 8AM to 4PM instead of 5PM), as minor overlap in some sessions was removed. The majority of sessions were deemed appropriate and retained, as were the graded curriculum with sessions building on each other and free time during the day for group activities. Small groups were rotated so that all residents had the opportunity to work with all their colleagues.

### Evaluation of ASK as a scholarly project

This was a mixed-methods design: Knowledge gains and retention were assessed using a standardized quiz and in the second year (2017), we undertook 2 focus groups.

### Participants

Participants included all members of PGY1 at the University of Manitoba from 2016-2018 (n=32). All wrote pre- and post-rotation quizzes. We added a comparison group of senior residents not exposed to ASK in 2018. Of 11 senior residents, 8 agreed to participate. For confidentiality, demographics were not collected.

### Study outcomes Quantitative

A quiz assessed knowledge before (time0, 2016-2018), immediately after (time1, 2016-2018), and 6 months after ASK (time2, 2017-2018) (see Appendix B for quiz). The quiz was mostly short-answers, though one scenario tested critical thinking in data interpretation. It was adapted from a national OSCE station from 2014. Assigned marks were reported as percentages. Tests were marked by a single author (CR) to ensure consistent scoring, well after ASK to avoid biasing evaluations.

Scholarly activity was assessed as the percentage of resident abstracts out of all abstracts at our local research day from 2014 to 2020 (range 58-90 per year). Typically, all abstracts submitted are accepted for presentation.

### Statistical analyses

Pre- and post-test scores were compared using paired and unpaired t-tests. Data are presented as mean and standard deviation (SD). A test for linear time trend was used to test for association with research productivity. A sample size analysis showed that 10 paired results were required for 80% power to detect a 10% difference from pre- to post-test. All analyses were performed in R version 4.0.3 with a two-sided p<0.05 considered statistically significant[20].

### Qualitative analysis

To assess resident expectations and satisfaction, pre- and post-rotation focus groups in the second year were facilitated by an author (DB) unaffiliated with the program to minimize bias. The pre-group was to explore expectations for ASK and experience and comfort in academic activities, such as teaching, feedback, biostatistics, QI and study design. The post-group was to determine if ASK met expectations, review comfort in the same academic areas, and suggest revisions. Focus groups were restricted to the 2017 cohort (n=11) to avoid contamination of future cohorts.

Focus groups were recorded, transcribed, and stripped of identification by DB before analysis by MB and DB using thematic analysis[21,22]. Differences were resolved by consensus.

## Results

Curriculum created in 2016 (Table 1) is now considered an essential component of PGY1 training. All residents were able to complete and pass the rotation. Minor modifications have occurred in the curriculum since the original, based on feedback from residents and preceptors and input from the ASK committee.

### Evaluation: Quantitative

All PGY1s participated in ASK (2016-2018) with 32 residents writing the tests at time0 and time1 (Table 2). The knowledge retention quiz was completed by 21 residents at time2 = 6 months. In 2018, eight senior non-ASK residents wrote the same test for comparison.

**Table 2.** Comparison of knowledge test scores.

|  | **Number of participants** | **Exam score**<br>**% Mean ± SD** | **P-value** |
| --- | --- | --- | --- |
| Pre-ASK (time0) | 32 | 52.6 ± 11.0% | -- |
| Immediately post-ASK (time1) | 32 | 80.2 ± 9.0% | <0.001 |
| 6 months post-ASK (time2) | 21 | 70.2 ± 12.0% | <0.001 (vs. time0) |
|  |  |  | <0.001 (vs. time1) |
| Non-ASK residents | 8 | 59.7 ± 18.7% | 0.02 (vs. time1) |
|  |  |  | 0.2 (vs. time2) |

### Knowledge Test

Compared to baseline, residents in the ASK demonstrated significantly increased knowledge immediately after the rotation (80.2 ±9.0% vs. 52.6 ± 11.0%, p<0.001). Although scores dropped somewhat after 6 months (70.2 ± 12.0%), they remained higher than baseline (p<0.001). Immediately after ASK, scores were higher than those of non-ASK senior residents (80.2 ±9.0%, vs. 59.7 ± 18.7%, p<0.02). Six months later, there was no significant difference between ASK PGY1 and senior non-ASK residents’ scores (70.2 ± 12.0% vs. 59.7 ± 18.7%, p=0.2).

### Resident Research Productivity

We noted an increase in the number of residents’ abstracts presented as a proportion of all abstracts at the local pediatric Research Institute research days, from 0% in 2014 to 8% in 2020 with a test for linear trend r=0.74, p = 0.01 (Figure 1).

**Fig1 legend:**
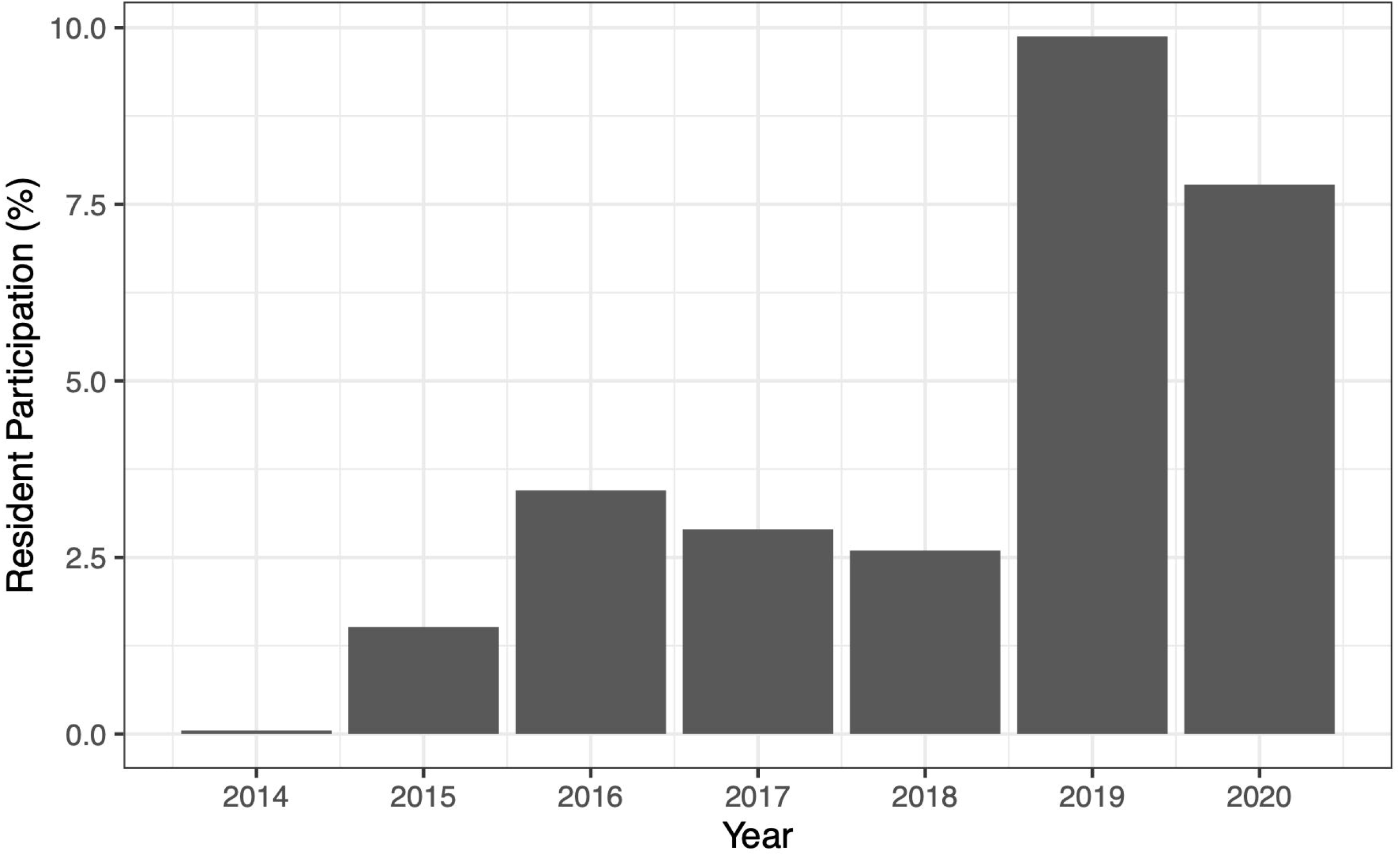
Percentage of abstracts authors by residents from 2014 to 2020 Percentage of abstracts authored by residents presented out of total number of abstracts presented annually at local pediatric Research Institute Day from 2014 to 2020. The first ASK rotation was held just at the annual autumnal research day in 2016.

### Evaluation: Qualitative

#### Pre-rotation focus group

The pre-rotation focus group identified residents’ goals that could be summarized as “theory to practice.” There were two major themes: Growth from learner to expert and developing skills in critical appraisal (Table 3).

**Table 3.** Pre- and post-rotation focus groups representative quotations.

|  | **Subthemes** | **Representative quotes** |
| --- | --- | --- |
| a. | School vs. real-world knowledge | *I think that now it seems more relevant. You're like, OK, this is why I need to know the stats because I need to critically appraise this article and it would be useful to know what the test is and why they are using it.*<br>*The only expectation was to learn how to do the tests for the exam and ... it wasn't incorporated into critical appraisal or actual real-world applications.* |
| b. | Starting research from scratch | *...all of the research projects that I have done, I started becoming involved in the data analysis stage, so all of the data was collected for me basically, so learning about the beginning parts of how to, like, collect participants, how to get approval, like all the ethics stuff, I have no idea about.*<br>*I feel that my experience with study design is minimal ... I don't know how I would, like, set up the study design and would definitely like guidance in that area.* |
| c. | Knowing where to get answers | *I feel like it is a realistic expectation for all of us to come to the end of this ... with the ability to ... know who to ask, know when we're on the wrong path*<br>*I am expecting to have more connections to people who are highly involved in research because ... I don't know those people. So knowing who to talk to when you want a specific type of help is hopefully going to be helpful.* |
| d. | Conflicted feelings about feedback | *I don't like getting negative feedback, it's hard for me to take negative feedback, but I like getting feedback ... I really like constructive feedback. I have had preceptors who were very good at saying, you know, "you could do this to work on this, you could do this" and just the way that they do it ... I don't take it as a negative thing. But there are other preceptors where ... I would get negative feedback and would be like, devastating just the way it was given. But ... I hate it when preceptors just say "you're doing great" because that's not useful.*<br>*I don't think anyone likes giving or receiving constructive criticism, but it's necessary for learning. With pats on the back like "keep up the good work" ... it doesn't help you at all. It's like, OK, cool, whereas if someone gives actual constructive feedback ... then you at least know what to work on.* |
| e. | Feedback recipients' openness | *I maybe am a little bit more hesitant to give constructive feedback, 'cause a lot of people say they want it [but] don't want to hear it*<br>*It sometimes doesn't really matter what level they are at, it's how willing they are to receive the feedback. So I just came off a rotation where the staff sat me down at the end and was like, ... "tell me everything..." and it felt like a really easy, open forum to give feedback ... And then, we also had a junior on that rotation who I think several of us tried* |
|  |  | *to give feedback and it wasn't really well received. So it's kind of the opposite situation to what I've been used to. Usually, I've been more comfortable giving feedback to juniors versus staff, but I think it comes down to how ready someone is to hear what you have to say* |
| f. | Giving constructive feedback | *I have liked it better when preceptors are like "this is what you do really well, and you could do this, this and this to be even better" ... I would like to be better at giving feedback so I can pass that on*<br>*I think when I give feedback ... I'm not that skilled at ... identifying what someone should be doing to work on those areas.* |
| g. | Strong foundation | *I think in medicine we are taught systematic, RCT ... but all of the other kinds of research have value and certain purpose. And I think through this process it will be good for everyone to be able to see when those things can be helpful, but also to then be able to read and understand what is that actually showing...* |
| h. | Comfort with statistics | *... you have this, like, basic framework and you kind of know whether something was a reasonable test to do for a question or whether it was significant, but actually doing it would be very difficult for me*<br>*I think we all probably have a basic grasp like ... we can, like, generally say, "ok, I sort of ... understand what this means" but I don't know if I would be able to come up with a summative sentence that says, like, this means this, because, like, the 'r' is this, the 'p' is this, the '95% CI' is this ...* |
| i. | Having a framework | *As a beginner, it really helps to have something that is step-by-step that kind of tells you all the points you are supposed to hit on, like, for critically appraising a paper.* |
| j. | Applying research to clinical practice | *... a resident has to know the difference between, like, this is significant statistically, but is it applicable to the patient or not? Not every p-value below 0.05 is significant for every patient.*<br>*What I'm not sure about is when ... you have a really poorly done RCT full of bias, sponsored by drug companies that are very influential in the actual study, and then you have a well done other type of study... How do you weigh those against each other? Is it still better 'cause it's a randomized control study but it's full of bias?* |

**Post-rotation focus group representative quotations**
|  | **Subthemes** | **Representative quotes** |
| --- | --- | --- |
| a. | Learning step-by-step | *You had your question about the progression and I think that was good. Like, we did the journal as a group which I think was helpful, to, like, bounce ideas off each other. But then later we had the opportunity to critique a journal on our own, which I think was important as well.* |
| b. | Increased confidence | *I might not be comfortable 100% of the time, but I feel like I have a better idea of resources to use when I am not sure, and kind of a foundation to fall back on.* |
| c. | Clinical relevance | *...the articles we were reading were SUPER clinically relevant to our practice ... there was a lot of gen[eneral] peds learning that we were able to do.* |
| d. | Improved feedback | *I do think the feedback given on the first two days is extremely different than the way feedback ... is given now. ... And even if people are still uncomfortable ... what you do and say will be better because of the experience some of us had here. ... Like, I do think the quality of the feedback and the specificity of it has, like, 100% improved.* |

##### Theme 1: Growth from learner to expert

Six subthemes are listed in Table 3 a-f with representative quotations. All agreed that although they may have previously covered ASK topics (“school knowledge”), they would approach them with a new lens. This included application to patient care and a deeper understanding that would allow them to apply information from the literature in practice (“real-world knowledge” a). This was particularly relevant as they considered their own scholarly projects. While most had previously participated in projects, few had undertaken an independent project (b). Although participants were hoping to gain confidence in knowing where to find answers to their questions (c), they were particularly interested in being better prepared for their own projects.

Participants were able to identify the tension between wanting feedback so they could improve and the difficulty receiving negative feedback (d). They were divided as to whether they were more comfortable giving feedback to people more junior vs more senior; however, most agreed that the recipients’ interest in receiving the feedback was an important factor in determining their comfort levels (e). They were more comfortable giving positive feedback than negative, so participants wanted to develop skills to ensure that their feedback would be concrete, actionable, and constructive (f).

##### Theme 2: Developing skills in Critical Appraisal

Four subthemes are listed with quotes in Table 3 (g-j). Participants agreed they were most comfortable with randomized control studies and systematic reviews and less comfortable with other studies (g). Everyone agreed they wanted to become more comfortable with biostatistics – both to be better able to appraise publications and to apply to their own future project (h). Most felt they still needed guidance when critically appraising a research paper (i). All felt inexperienced in applying research to clinical medicine (j).

#### Post-rotation focus group

Table 3 shows the four subthemes (a-d). Feedback was overwhelmingly positive; ASK met expectations and residents appreciated the time to learn the material gradually while practicing (a). They felt more confident when appraising scientific literature (b) and appreciated the articles’ relevance to pediatrics (c). After a didactic session in week 1, residents practiced daily peer-to-peer feedback, and all felt they had improved in their ability to give concrete feedback (d). They also made suggestions for future improvements.

An additional benefit of the rotation was increased camaraderie, observed by the focus group facilitator and in a more animated transcript. Post-rotation, one participant noted that they were “like siblings now!”

## Discussion

We were pleased to have been able to develop, implement, and evaluate the new ASK curriculum. It expanded on earlier longitudinal teaching. to support scholarly activity and now encompasses a robust approach to teach additional academic skills, such as QI, teaching multilevel learners, and becoming a better lifelong learner. Our evaluation strongly suggests that knowledge is enhanced and retained in PGY1 and now matches that of senior residents, who were not exposed to ASK. Moreover, PGY1s welcome this rotation. Additionally, it would appear to support scholarly activities through increased participation in the annual Childhood Research Days (2014-2020). ASK is now a cornerstone of CBD as the residents ‘transition to discipline’ (i.e. pediatrics).

While most previous studies assessing the impact of programs supporting research skills have not involved pediatric residents, their results are nonetheless generalizable [6–9,23,24]. They included research curricula, introduction to potential supervisors, or appointment of an experienced research coordinator. Generally, all found increased knowledge in pre-post exams[5,7,8,23]. Others report resident satisfaction, increased participation in local or external conferences, and increased publications[6,9,11,12,25].

This study adds to the literature by confirming knowledge retention 6 months after the rotation, with significantly higher test scores 6 months after the rotation compared to the pre-test. This comparison group of senior residents would have been expected to gain similar knowledge from academic sessions throughout their residency and through completion of a scholarly project. Our study demonstrates that tests scores of first year ASK participants 6 months after the rotation were not significantly different to those of the senior residents, though knowledge may be acquired more efficiently in ASK. We cannot speculate how residents who have undertaken ASK will perform on this quiz as senior trainees. We believe that their knowledge will be reinforced by other academic activities, such as journal clubs, academic half-days, and completion of research projects.

There are few other qualitative assessments of such curricula in the literature [8]. The pre-rotation focus group demonstrated that expectations were similar to those anticipated during curriculum development. These themes have been broadly categorized as ‘theory to practice’. This fits with constructs described by Noble *et al*[26]. Many of these concepts have been studied in undergraduate or medical school, and ASK reinforced conceptual knowledge of EBM – adding in pediatric specific examples (by reviewing seminal pediatric articles) – to build conceptual and pediatric care knowledge.

Having anticipated their desire for additional conceptual knowledge, it is perhaps not surprising that feedback in the post-rotation was positive. We also embraced increased procedural knowledge with mandatory biostatistics assignments, individual presentation of journal articles, rapid literature searches, and discussion of pediatric topics to peers in a safe environment. These early procedural steps will be reinforced throughout their training, particularly as they work on their mandatory scholarly projects. We believe ASK increases confidence early in training, which is reinforced by the other supports in place for scholarly projects[5,25,26].

As previously reported, resident feedback supports setting aside protected non-clinical time allowing for the development of skills sequentially without interruption[8,27]. Given positive reactions to articles that were relevant to pediatrics, other residency programs may also want to focus on articles from their own clinical specialties to ensure resident engagement and gains in general knowledge through article critiques.

Unlike the post-NERD protected time in Canadian Emergency Room program, where trainees in the post-course focus group felt conflicted between scholarly enrichment and lifelong learning, our post-course feedback did not identify this issue [8]. Our trainees understood that ASK was not just focussed on scholarly projects, but provided a foundation for lifelong learning and better patient care. Approximately 50 research-active faculty preceptors participated in the rotation, representing local university- and hospital-affiliated researchers[26]. The rotation may also facilitate faculty-resident research collaboration as observed in other studies [8]. We were also fortunate to have support from both the Department of Pediatrics and the Research Institute, who generously liberated residents and hospital staff from clinical duties and research staff from other teaching and research responsibilities.

Despite initial discomfort with peer-to-peer feedback, resident distress was much reduced by the end. We followed the steps outlined by de la Cruz *et al*, with an initial preparatory session deliberately offered in the first week and standard evaluation forms used by peers and staff-preceptors[28]. Both peers and faculty provided daily verbal and written feedback for individual presentations, and a marked change in attitudes was noted in the pre-post focus groups. The residents appreciated that peer feedback - especially with more senior colleagues - can be challenging. Nonetheless, they appeared to understand that these were important professionalism skills[29].

We were also pleased to note increased scholarly activity with ASK, as reported with comparable rotations[9,24,27]. Previously, the most frequently identified major barrier to completing a scholarly project was lack of time[15,30]. However, the lack of a research curriculum, statistical support, and funding have also been identified[30]. Our rotation has likely helped address some of these barriers, which may explain the increased scholarly activity from our residents since the introduction of ASK. Although establishing a causal relationship between this rotation and scholarly productivity is difficult.

The strengths of our study include 3 years of data, assessment of both longer-term knowledge retention and research productivity, and inclusion of a comparison group. See appendices C-E for materials used in course, feedback forms and grids for resident feedback.

This study is limited to small sample sizes within a single residency program. We did not have access to a validated quiz and adapted a previously used OSCE tool. Although the quiz was not modified over time; residents were not warned in advance. With a 6-month gap before the post-test, we cannot exclude learning effects. A better pre- post-comparison might have been provided by assessing journal critiques as a broader measure of academic skills. Possible contamination from the 2017 focus group precluded similar evaluations in later years, and we do not know if we reached data saturation.

## Conclusion

A 4-week non-clinical rotation dedicated to academic skills - including critical appraisal, teaching, QI, research evaluation, biostatistics, and professionalism – was associated with increased knowledge retained over 6 months and increased resident scholarly activity. Our description should allow other programs to incorporate aspects into their own academic training. Residents appreciated the time to learn the material, especially since materials were pediatric-specific. Our study was novel in that it looked at broader academic skills and longer term retention, included a control group of senior residents, and utilized mixed-methods to better explore expectations and areas for improvement.

## Data Availability

All data produced in the present study are available upon reasonable request to the authors

## Declarations

Compliance with Ethical Standards

None of the authors declare any conflicts of interest.

Ethics approval was obtained from the University of Manitoba Research Ethics Board (HS20909: H2016:219- Date August 27, 2016). Informed consent to participate in the study was obtained from residents by a research assistant not affiliated with the program

## Appendices

A. Goals and Objectives

B. Pre- and post-rotation quiz

C. Materials used in course

D. Feedback forms

E. Grid for resident feedback about individual sessions

## References

[1] CanMEDS Framework□:: The Royal College of Physicians and Surgeons of Canada n.d. https://www.royalcollege.ca/rcsite/canmeds/canmeds-framework-e (accessed February 8, 2021).

[2] Accreditation Council for Graduate Medical Education. Common Program Requirements 2020. https://www.acgme.org/What-We-Do/Accreditation/Common-Program-Requirements (accessed February 8, 2021).

[3] Hatala R, Guyatt G. Evaluating the teaching of evidence-based medicine. JAMA 2002;288:1110–2. https://doi.org/10.1001/jama.288.9.1110.

[4] Wood E, Kronick JB, Association of Medical School Pediatric Department Chairs, Inc. A pediatric residency research curriculum. J Pediatr 2008;153:153–4, 154.e1-4. https://doi.org/10.1016/j.jpeds.2008.02.026.

[5] Thom DH, Haugen J, Sommers PS, Lovett P. Description and evaluation of an EBM curriculum using a block rotation. BMC Med Educ 2004;4:19. https://doi.org/10.1186/1472-6920-4-19.

[6] Ruiz J, Wallace EL, Miller DP, Loeser RF, Miles M, Dubose TD, et al. A comprehensive 3-year internal medicine residency research curriculum. Am J Med 2011;124:469–73. https://doi.org/10.1016/j.amjmed.2011.01.006.

[7] Farrokhyar F, Amin N, Dath D, Bhandari M, Kelly S, Kolkin AM, et al. Impact of the Surgical Research Methodology Program on surgical residents’ research profiles. J Surg Educ 2014;71:513–20. https://doi.org/10.1016/j.jsurg.2014.01.012.

[8] Abu-Laban RB, Jarvis-Selinger S, Newton L, Chung B. Implementation and evaluation of a novel research education rotation for Royal College of Physicians and Surgeons emergency medicine residents. CJEM 2013;15:233–6. https://doi.org/10.2310/8000.2013.130941.

[9] Roth DE, Chan M-K, Vohra S. Initial successes and challenges in the development of a pediatric resident research curriculum. J Pediatr 2006;149:149–50. https://doi.org/10.1016/j.jpeds.2006.05.001.

[10] Horsley T, Hyde C, Santesso N, Parkes J, Milne R, Stewart R. Teaching critical appraisal skills in healthcare settings. Cochrane Database Syst Rev 2011:CD001270. https://doi.org/10.1002/14651858.CD001270.pub2.

[11] Wood W, McCollum J, Kukreja P, Vetter IL, Morgan CJ, Hossein Zadeh Maleki A, et al. Graduate medical education scholarly activities initiatives: a systematic review and meta-analysis. BMC Med Educ 2018;18:318. https://doi.org/10.1186/s12909-018-1407-8.

[12] Stevenson MD, Smigielski EM, Naifeh MM, Abramson EL, Todd C, Li S-TT. Increasing Scholarly Activity Productivity During Residency: A Systematic Review. Acad Med 2017;92:250–66. https://doi.org/10.1097/ACM.0000000000001169.

[13] Kern, DE, Thomas, PA, Hughes, MT. Curriculum Development for Medical Education: A Six-Step Approach. Second. Baltimore: Johns Hopkins University Press; 2009.

[14] Pediatric Resident Research. Pediatric Resident Research n.d. https://pedresresearch.ca (accessed March 1, 2022).

[15] Pound CM, Robinson J, Giglia L, Rodd C, Sharma A, Chafe R, et al. Scholarly training objectives and requirements for paediatric residents in Canada. Paediatr Child Health 2019;24:76–80. https://doi.org/10.1093/pch/pxy070.

[16] Competence by Design□:: The Royal College of Physicians and Surgeons of Canada n.d. https://www.royalcollege.ca/rcsite/cbd/competence-by-design-cbd-e (accessed March 1, 2022).

[17] Harvey, B, Lang E, Frank J. The Research Guide: A primer for residents, other health care trainees, and practitioners. First. The Royal College of Physicians and Surgeons of Canada.; 2011.

[18] IHI Open School Home | IHI - Institute for Healthcare Improvement n.d. http://www.ihi.org:80/education/IHIOpenSchool/Pages/default.aspx (accessed March 1, 2022).

[19] TCPS 2: CORE-2022 n.d. https://tcps2core.ca/welcome (accessed March 1, 2022).

[20] R: The R Project for Statistical Computing n.d. https://www.r-project.org/ (accessed February 9, 2021).

[21] Breen R. A Practical Guide to Focus-Group Research. Journal of Geography in Higher Education 2006;30:463–75.

[22] Maguire M, Delahut B. Doing a Thematic Analysis: A Practical, Step-by-Step Guide for Learning and Teaching Scholars. All Ireland Journal of Teaching and Learning in Higher Education n.d.;9:3351–33514.

[23] Löwe B, Hartmann M, Wild B, Nikendei C, Kroenke K, Niehoff D, et al. Effectiveness of a 1-year resident training program in clinical research: a controlled before-and-after study. J Gen Intern Med 2008;23:122–8. https://doi.org/10.1007/s11606-007-0397-8.

[24] Rothberg MB, Kleppel R, Friderici JL, Hinchey K. Implementing a resident research program to overcome barriers to resident research. Acad Med 2014;89:1133–9. https://doi.org/10.1097/ACM.0000000000000281.

[25] Mills LS, Steiner AZ, Rodman AM, Donnell CL, Steiner MJ. Trainee participation in an annual research day is associated with future publications. Teach Learn Med 2011;23:62–7. https://doi.org/10.1080/10401334.2011.536895.

[26] Noble C, Billett SR, Phang DTY, Sharma S, Hashem F, Rogers GD. Supporting Resident Research Learning in the Workplace: A Rapid Realist Review. Acad Med 2018;93:1732–40. https://doi.org/10.1097/ACM.0000000000002416.

[27] Vinci RJ, Bauchner H, Finkelstein J, Newby PK, Muret-Wagstaff S, Lovejoy FH. Research during pediatric residency training: outcome of a senior resident block rotation. Pediatrics 2009;124:1126–34. https://doi.org/10.1542/peds.2008-3700.

[28] de la Cruz MSD, Kopec MT, Wimsatt LA. Resident Perceptions of Giving and Receiving Peer-to-Peer Feedback. J Grad Med Educ 2015;7:208–13. https://doi.org/10.4300/JGME-D-14-00388.1.

[29] Çoruh B, Kritek PA. Implementation of a Coaching Program for Peer Feedback on Large-Group Teaching. Ann Am Thorac Soc 2017;14:601–3. https://doi.org/10.1513/AnnalsATS.201611-936LE.

[30] Abramson EL, Naifeh MM, Stevenson MD, Mauer E, Hammad HT, Gerber LM, et al. Scholarly Activity Training During Residency: Are We Hitting the Mark? A National Assessment of Pediatric Residents. Acad Pediatr 2018;18:542–9. https://doi.org/10.1016/j.acap.2018.02.002.

