## Appendix A for "Creation of novel pediatric academic curriculum and its evaluation using mixed methods"

**ROTATION GOALS & OBJECTIVES**

**Academic skills and knowledge rotation**

**GENERAL**

The Academic skills and knowledge (ASK) rotation is a mandatory pediatric PGY1 residency rotation designed to provide pediatric residents with tools and the opportunity to practice of research, teaching and communication skills. The rotation will occur yearly in period 5 and all PGY1 residents complete the rotation concurrently. Residents are not scheduled for call during the weekdays as it is expected that attendance will be 100%. Residents will still be expected to take call one Friday and one Saturday during the 4-week rotation. Preparatory work will be assigned periodically throughout the rotation in order to facilitate the small group teaching environment. There will be a variety of teachers during the rotation and specific tasks will be completed by the resident. The MRA and the ITAR will be completed by Dr. Celia Rodd and these assessments will be done face to face. All sessions will be held in room 474A in the Chown building. This large classroom provides space for breakout groups and up-to-date audio-visual equipment.

**RESPONSIBILITIES**

Faculty

1. Dr. Celia Rodd will review the expectations, goals and objectives of the rotation on the first day of the rotation as well as the relationship to the CanMEDS competencies.
2. Teaching faculty are expected to provide feedback to residents in real time and provide critiques of the expected tasks. This will provide continuous improvement opportunities to the residents.
3. Faculty must complete the written evaluation AND give face-to-face feedback to the resident at the middle and end of each rotation. Written evaluations are to be completed on VENTIS within 21 business days of the close of the rotations.

Resident

1. The resident must thoroughly read this curriculum prior to the start of the rotation.
2. **Teaching development program 1(TDP1) and Teaching development program 2 (TDP2), online core curriculum modules, must be completed prior to the beginning of the rotation. Access to these modules is found on the website listed below:**

u**manitoba**.ca/faculties/medicine/education/pgme/**core**_**curriculum**.html

3. The resident should review this curriculum with the supervising faculty on the first day of the rotation. Attendance is mandatory and 100% expected. Resident is expected to complete all preparatory work and individual assessment opportunities assigned (e.g. preparing a poster, preparing a critically appraised topic (CAT).
4. The resident must review his/her performance with the supervising faculty informally throughout the rotation and formally at the middle and end of the rotation.

An interim evaluation is mandatory midway through each rotation as per University requirements. A final evaluation is provided at the end of the rotation. The resident is also evaluated by each service attending with whom he or she has been on-service.

Academic credit for this rotation will be granted upon the completion of the following three items: resident evaluation by the preceptor(s) and resident’s online evaluations of both the rotation and the preceptor(s) within 21 business days of the end of the rotation.

**SCHEDULE:**

**Week 1**

| Thursday | Friday | Mon | Tues | Wed |
| --- | --- | --- | --- | --- |
| Grand rounds | 8-830 Literature search follow-up – review the bibliographies that were created the day before  (30 min) | Study design: descriptive vs. interventional, RCT, case-control, prospective observational, cross-sectional observational, administrative, retrospective (chart review)-2h | Critical appraisal by study type- case control, retrospective, cross-sectional, cohort, RCTs | 8-830 Present on authorship roles, retention and ethics of recruitment |
|  |  |  |  | 830- 930Facilitating small group teaching |
|  | Statistics: Predictors, Exposures, and variable types i.e.  descriptive stats, t tests and chi-squared tests |  |  |  |
| Introduction |  | Limitations, confounders, bias | 1^st^ journal club – only Intro and Methods | Practice leading a small group—(we provide topics) |
| Asking clinical and research questions | How to give effective feedback + receive | Find examples of different study designs in pediatric journals | Each student then presents their critique + feedback (50% pre WIP) | Feedback |
| CHRIM rounds |  |  | WIP | Academic half-day |
| Tips on how present—journal club, CAT, slides |  | Finding a supervisor and resident-supervisor relationship expectations | Each student then presents their critique + feedback (50% post WIP) |  |
| Library search strategies | Practical lab-basic stats  (use text commands and buttons) | Research ethics – patient confidentiality and safety | 30 min short question—search and learn |  |
| Learning how to use Zotero  - Create a bibliography  (we will provide the question) |  |  |  |  |
|  |  | Funding for scholarly projects | 5 min presentations of 30 min search and learn |  |
|  |  | QI- 1^st^ hour | Feedback | 345-445  Senior resident –how select project |
| Life long learning |  |  |  |  |
|  |  |  | Speed dating |  |

**Targeted material and tasks to accomplish during week 1:**

1. Conduct a literature search.
2. Develop a bibliography.
3. Perform a rapid focused literature search a topic and synthesize the data.
4. Critique a journal with basic stats and study design knowledge.
5. Learn steps on how to lead small group learning and teaching.

**Week 2**

| Thursday | Friday | Mon | Tuesday | Wed |
| --- | --- | --- | --- | --- |
| Grand Rounds | 8-830 review homework | 8-830 review homework | Elements of a scholarly project proposal + timelines | Ethics tutorial |
|  |  | Budgeting & study costs |  |  |
|  | Consents – theory and practice  1 h theory |  |  |  |
| Stats - sample size (theory +manual calculations)2h |  |  |  |  |
| QI 2^nd^ hour | How to ask consent/ permission to contact | How to synthesize or prepare a CAT: research tips, presentation tips, and assigned questions | QI – 2h | The art of submitting ethics, IMPACT, FAAF, forms and using a CRF |
|  | How to teach medical students | Residents need to find 5 manuscripts for a CAT |  | Research cultural sensitivities |
|  |  |  |  | Time management skills |
| CHRIM rounds |  |  | WIPs |  |
| Data entry + handling: data cleaning, wide vs. long, variables types, data dictionaries, use of databases options – Excel, Access , REDCap, importing + exporting data | Additional types of scholarly endeavours and study designs:  education, QI, qualititative, systematic reviews, administrative | How to teach effectively in 5 min or less | Review assigned journal article- 1 h to cover Intro, Methods and Results  (2 -3residents per group)  several articles – same theme | Academic half-day |
|  | Timeline for scholarly projects | Practice teaching in 5 min sessions | Teams present journal articles @ 30 min Critique and discussion  2.5h |  |
|  |  | QI – hours 3 and 4 | Provide the 5 articles they have selected for the CAT  (can be done earlier in the week, too) |  |
| More speed dating | Administrative  database analyses |  |  |  |
|  | Timeline for project | SPEED DATING |  |  |

**Targeted material and tasks to accomplish during week 2:**

1. Undertake essential statistical analyses.
2. Understand the elements of ethics in research by creating a informed consent form.
3. Understand the elements of teaching medical students (either brief – 5min teaching moments or longer more formal sessions).
4. Undertake a 5 min session (with feedback given and received – reinforcing how to provide feedback).
5. Prepare and present a critically appraised topic (CAT).
6. Undertake a more extensive journal club presentation.

**Week 3**

| Thurs | Friday | Mon | Tues | Wed |
| --- | --- | --- | --- | --- |
| Grand Rounds | Diagnostic studies | Discuss proposals with committee as a group | Time to review and discuss topics to date (30min) | 1^st^ team project 30 in prep time- QI or educational question-  15 min per group to present and defend approach  how to tackle this topic   - what is the best design - pros and cons of other designs |
| Stats: regression theory |  |  |  |  |
| Regression Lab- hands on | The importance of pre-assessing the utility of the tests that we order | Work on proposal | Present journal articles- -focus on Discussion of previously discussed article (1h to prepare)-by group  -include a brief lit search to look at other articles on these topics (team) |  |
|  | ½ present their CAT and feedback  15 + 10 min |  | Present articles+ feedback |  |
| CHRIM rounds |  |  | WIP |  |
| Questionnaire- design and validation  - types of responses, proxy measures | ½ present their CAT and feedback  15 + 10 min per residents | How to write a manuscript | Tips to making presentations- oral and poster | Academic half-day |
|  |  | Advocacy | Poster tour |  |
| How to develop research questions- BYI- bring your idea  (Small groups each with a facilitator) |  | Development of a portfolio | Discuss proposals with committee as a group- have 2 or 3 residents meet with a preceptor |  |
|  | Work on 2 page scholarly project proposals |  |  |  |

**Targeted material and tasks to accomplish during week 3**

1. Complete and provide us with a copy of the TCPS ethics online certificate.
2. Know how to prepare an ethics submission.
3. Draft a scholarly project protocol.
4. Perform and interpret regression analyses.
5. Know how to prepare a questionnaire.
6. Know how to initiate a group project.

**Week 4**

| Thurs | Fri- Nov 11, 2016 | Mon | Tues | Wed |
| --- | --- | --- | --- | --- |
| Review existing CVs- and work on them | Work on journal club article | Additional training opportunities—CIP, MSc | 30 min to ‘search and learn’ | The other half presents their journal club articles |
| Authorship |  | 2^nd^ team competition—scholarly project | Present their search and learn topic 5 min each |  |
|  |  |  | KT | Post course survey + how to improve = 1h |
|  |  |  |  | LUNCH |
| CHRIM rounds |  |  | WIP |  |
| Interpreting data correctly- provide a set of tables and figures |  | Presentation of CAT poster + feedback | Present journal articles in teams (50%of the teams) |  |
| Communicating to the public |  |  | Work on personal or team proposals+ meet with supervisors |  |
| Using CAT topic- prepare an abstract + prepare a poster |  | Work on proposals+ plus can include supervisors | Open house with supervisors |  |

**Targeted material and tasks to accomplish during week 4**

1. Develop a protocol or an outline of one.
2. Learn how to prepare clear and concise presentations and present their protocol.
3. Translate detailed medical knowledge into information accessible to families.
4. Find a supervisor.
5. Further develop their own CV.
6. Present an additional journal club at the level that is expected at Pediatric J Club sessions.
7. Prepare a poster of their protocol.

**OBJECTIVES**

Objectives for this rotation are listed below in CanMEDS format. Objectives noted in **bold** are specific to this rotation; other items are more universal and may apply to every resident rotation. Many of these common items are reflected on the Royal College Final In-Training Evaluation Report (FITER). Resident evaluations will center on all items listed, which will be assessed in context of this particular rotation and post-graduate year of training.

**THE PEDIATRICIAN AS MEDICAL EXPERT**

1. **Develop a basic understanding of statistics required for practice of evidenced based medicine (including but not limited to t tests, p values, Odds ratio's, means, standard deviations, variables, type 1 and 2 errors etc.)**
2. **Develop an understanding of research study designs including but not limited to case control, cross-sectional, cohort, RCTs, retrospective, descriptive etc.**
3. **Demonstrate development of a clinical or QI (quality improvement) question, a CAT (critically appraised topic), a CV, an Ethics proposal, a written consent, a journal club article analysis, an abstract, data entry and data analysis, a research protocol, and a research manuscript.**
4. **Develop an approach to time management.**
5. **Demonstrate an approach to giving feedback, teaching medical students and presenting a short topic.**
6. **Become familiar with the elements of informed consent including adequacy of consent, complications of multilingual consent and types of consent (assent, implied etc.)**
7. **Demonstrate knowledge of library search strategies and how to develop a bibliography.**

**THE PEDIATRICIAN AS COMMUNICATOR**

1. **Present oral reports of CATs, journal club article analyses, abstracts, ethics proposals in an accurate, complete and organized fashion.**
2. **Convey relevant and accurate information and explanations to others while delivering a small group teaching session, role playing request for consent and medical student teaching, presenting a CAT, journal club article etc.**
3. **Prepare written documentation that is accurate and organized such as but not limited to 2 page summaries, ethics proposals, questionnaires, C.V.s, scholarly project protocols etc.**
4. **Elicit and synthesize relevant information and perspectives of others when discussing CATs, journal club presentations, WIPs, teaching presentations etc.**
5. **Learn the need to address diversity and differences and the impact this has on research decision-making and effective communication.**

**THE PEDIATRICIAN AS COLLABORATOR**

1. **Collaborate respectfully and effectively with other trainees and faculty while role playing, sharing tasks of critically appraised topics etc.**
2. Delegate tasks/roles appropriately AND when indicated

**THE PEDIATRICIAN AS LEADER**

1. **Learn the need for appropriate and cost-effective use of all forms of pediatric health care resources including laboratory investigations, imaging etc.**
2. **Use information technology to optimize research including search resources, data entry options (RedCAP/excel), statistical software etc.**
3. **Exhibit appropriate time management skills including being prepared for daily lessons with preparatory work done, arriving on time and participating in sessions.**
4. **Understand the need to be a leader in quality improvement.**
5. **Demonstrate knowledge of the Research Ethics board and its role in child health research**

**THE PEDIATRICIAN AS HEALTH ADVOCATE**

1. **Develop an approach to advocate for children with the media and at community and government levels.**

**THE PEDIATRICIAN AS SCHOLAR**

1. **Evaluate information and its sources critically and apply this appropriately to the research question/topic.**
2. **Effectively presents background information and conclusions relating to a question to fellow trainees in the form of a journal club article, WIP, Abstract, small group discussion etc.**
3. **Maintain and enhance professional activities through ongoing learning.**
4. **Perform critical and accurate self-evaluation of one’s performance. Recognize gaps in knowledge and expertise and develop strategies for improvement.**
5. **Facilitate the learning of other trainees and faculty.**
6. **Seek out and receive feedback well with a goal of incorporating constructive feedback into scholarly activities.**
7. **Give constructive feedback to teachers and students.**
8. **Contribute to the creation, dissemination, application and translation of new medical knowledge in the form of WIPs, journal club presentations, abstract presentation, scholarly project presentation etc.**

**THE PEDIATRICIAN AS PROFESSIONAL**

1. Demonstrate personal and professional attitudes consistent with a general consultant pediatrician, including honesty, integrity, commitment, compassion, empathy, respect and altruism.
2. Demonstrate respect for others and diversity.
3. Demonstrate reliability, responsibility and conscientiousness with meeting deadlines, being punctual, completing assigned duties and fulfilling commitments.
4. Demonstrate self-awareness; seek advice when necessary and accept advice graciously.
5. Show motivation and ability to learn.
6. **Recognize ethical issues encountered in research.**
7. Demonstrate recognition of the importance of work/life balance for practice sustainability.

**SPECIFIC TASKS:**

**Residents will be expected to keep a portfolio of specific tasks. These will be evaluated and include:**

1. **Oral presentation of journal article.**
2. **CAT (critically appraised topic and poster presentation).**
3. **Bibliography.**
4. **TCPS ethics certificate.**
5. **C.V**
6. **Two-page protocol**
7. **Statistics 1 submission**
8. **Statistics 2 submission**
9. **Regression outputs**
10. **Quality improvement certificate**
