## Appendix B for "Creation of novel pediatric academic curriculum and its evaluation using mixed methods"

ASK 2020

This is a test to evaluate your knowledge of some aspects of academic skills.

Please use the 30 min allotted to complete this.

This does not count towards your assessment in this rotation.

The following study appeared in *J Pediatr 2015: 167, 253-9*

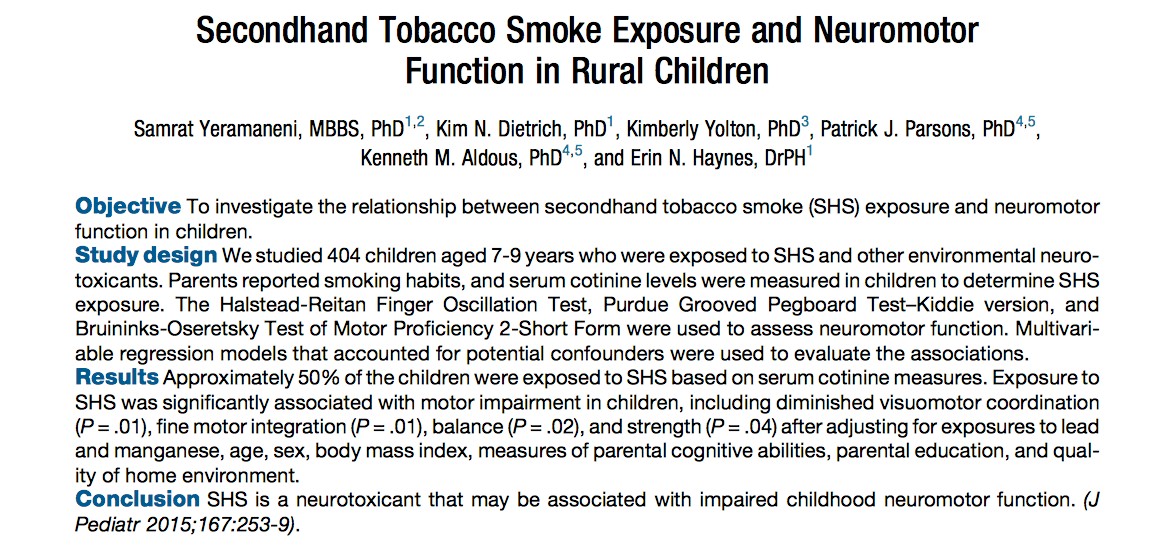

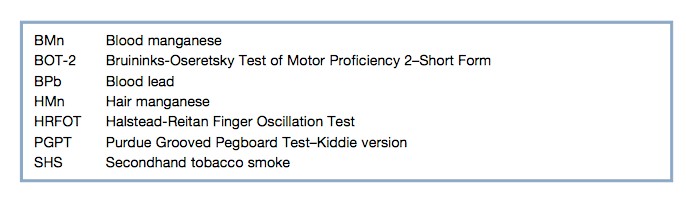

1) Identify the primary exposure variable and the primary outcome variable(s)

(1 point):

_____________________________________________________________________________________________

2) This is a table of Pearson correlation coefficients (r) between various exposure and outcome variables, also known as a correlation matrix. Interpret/explain the meaning of the Pearson r (1 point). Identify two limitations (2 points) of this measure

Table II. Bivariate associations (Pearson correlation, *r*) between logarithms of serum cotinine, BPb, BMn, and HMn levels and children’s neuromotor performance

|  | **Serum cotinine** | **BPb** | **BMn** | **HMn** | **Dominant hand HRFOT** | **Nondominant hand HRFOT** | **Dominant hand PGPT** | **Nondominant hand PGPT** | **BOT-2** |
| --- | --- | --- | --- | --- | --- | --- | --- | --- | --- |
| **Serum cotinine** |  |  |  |  |  |  |  |  |  |
| **BPd** | 0.35* |  |  |  |  |  |  |  |  |
| **BMn** | -0.09 | -0.12† |  |  |  |  |  |  |  |
| **HMn** | 0.25* | 0.22‡ | 0.001 |  |  |  |  |  |  |
| **Dominant hand HRFOT** | -0.14† | -0.18§ | 0.01 | -0.10 |  |  |  |  |  |
| **Nondominant hand HRFOT** | -0.19‡ | -0.21‡ | -0.02 | -0.03 | 0.69* |  |  |  |  |
| **Dominant hand PGPT** | 0.20‡ | 0.10 | 00.04 | 0.07 | -0.26* | 0.25* |  |  |  |
| **Nondominant hand PGPT** | 0.20‡ | 0.14† | 0.03 | 0.04 | -0.33* | -0.36* | 0.64* |  |  |
| **BOT-2** | -0.22* | -0.02 | -0.03 | -0.03 | 0.22‡ | 0.24* | -0.25* | -0.26* |  |

**P*< .0001, † *P*< 0.05, ‡ *P*< .001, § *P*< .01

This table contains the results of multivariate linear regression where the outcome variables are various neuromotor subscores (for simplicity, we’ve left only two sets visible). Serum cotinine is measured in ng/ml, age in years, and the reference level for sex (a 0/1 categorical variable) was set as female = 0.

3) Explain the meaning of the betas (partial regression coefficients) (1 point).

Table III. Multivariable associations between logarithm serum cotinine levels and children’s neuromotor performance

| **Neuromotor outcomes** | **Explanatory variable** | **ß (95%CI)** | ***P* value** | **Adjusted *R^2^*** |
| --- | --- | --- | --- | --- |
| Dominant hand HRFOT | Serum cotinine  Age  Sex*  BPb  HMn | -0.19 (-0.54 to 0.14)  2.52 (1.81 to 3.24)  1.61 (0.30 to 2.92)  -1.50 (-3.03 to 0.03)  -0.49 (-1.27 to 0.28) | .25  <0.001  .02  .05  .21 | 0.18 |
| BOT-2, Total motor composite score | Serum cotinine  BMI  Barratt’s Education  BPb  BMn (linear)  BMn (quadratic) | -0.64 (-1.13 to -0.16)  -0.44 (-0.69 to -0.20)  0.33 (-0.05 to 0.71)  1.18 (-0.71 to 3.07)  69.61 (20.80 to 118.43)  -15.63 (-26.41 to -4.85) | .009  .003  .09  .22  .005  .004 | 0.15 |

*Reference group is female

Explain the meaning of the 95% confidence intervals (1 point):

_________________________________________________________________________________

Explain the meaning of the p-­‐values (1 point):

_______________________________________________________________________

4) Explain the difference between statistical significance and clinical significance (1 point)?

5) Does this study provide evidence in support of a causal association between household exposure to second-­‐hand smoke and neuromotor outcomes (1 point):

________________________________________________________________________________

6) What is a confounder (1 point)?

_______________________________________________________________________________

Explain how the 23.3% rate of maternal smoking throughout pregnancy (Table 1, not shown) is a potential confounder (1 point)?

________________________________________________________________________________

7) Name 3 features of a solid research supervisor? (3 marks)

______________________________________________________________________________
_______________________________________________________________________________

______________________________________________________________________________

8) Name the kind of variable these are? (4 marks)

Age:___________________________________

Sex:____________________________________

Hypertension (mild/moderate/severe)____________________________

Race/ethnicity____________________________

9) Name 3 elements of an informed consent process (3)

______________________________________________________________________________________________________________________________________________________________________________________________________

10)

Match the phrase with its definition (2 points)

The proportion of patients who test positive actually have the disease

Negative predictive value

It is the probability of the subjects with a negative screening test do not have the disease

Sensitivity

The proportion of the nondiseased people who are correctly identified as negative by the test

Positive predictive value

The proportion of the diseased people who are correctly identified as positive by the test

Specificity

11) For each study design list 1 positive attribute and 1 draw back (6 points)

a) case-control

______________________________________________________________________________________________________________________________________________________________________________________________________

b) cohort

______________________________________________________________________________________________________________________________________________________________________________________________________

c) qualitative

______________________________________________________________________________________________________________________________________________________________________________________________________

12) Quality Improvement (QI) work describes a PDSA cycle (2 points)

What do the letters P-D-S-A stand for

13) When calculating a sample size for a project-what information do you need for a study comparing age of onset of STIs in adolescents vs. young adults? (3 points)

___________________________________________________________________________________________________

___________________________________________________________________________________________________

14) When being a small group facilitator, you are faced with a student who never answers any questions. How can you work to involve them in answering? (1 point)

________________________________________________________________________________________

15) In a manuscript, the Methods section contains important information

List 3 of the standard types of information (or subsection headings) for a Methods section (3 points)

Total 38 points
