## Appendix C for "Creation of novel pediatric academic curriculum and its evaluation using mixed methods"

Materials used in course

1. Computer and A-V projector
2. Black or white board
3. Flip chart
4. 2 simulation mannequins plus simulation assistants to oversee multilevel learner teaching (2 hours in the course)
5. QI team to help with simulation practice scenario exploring optimizing patient flow from admission to discharge; this involved 6 additional staff, some mock patient information and all residents (duration 2 hours)
