## Appendix D for "Creation of novel pediatric academic curriculum and its evaluation using mixed methods"

Presentation (circle the appropriate one)

A) Search and learn and presentation
B) CAT
C) Small group teaching
D) Team competition 1 or 2
E) Journal club—team vs. individual
F) CV
G) Stats homework 1 2 3 4
H) PDSA ideas and concepts
I) Abstract creation

Name of preceptors or residents providing feedback ________________________

**Topic ________________	Resident presenting________________	Date_______**

Communication style

| Significantly below expectations | Slightly below expectations | Meets expectations | Slightly above expectations | Significantly above expectations |
| --- | --- | --- | --- | --- |

Global assessment

| Significantly below expectations | Slightly below expectations | Meets expectations | Slightly above expectations | Significantly above expectations |
| --- | --- | --- | --- | --- |

List 2 strengths

List 2 areas for improvement
