## Appendix E for "Creation of novel pediatric academic curriculum and its evaluation using mixed methods"

| **Date** | **Time** | **Title** | **Facilitator** | **Keep** | **Modify** | **Delete** | **Comments** |
| --- | --- | --- | --- | --- | --- | --- | --- |
| Sept 24  Thurs |  |  |  |  |  |  |  |
|  | 0930-1050 | Introduction |  |  |  |  |  |
|  | 1100- 1150 | ASKING a research Question |  |  |  |  |  |
|  | 1315-1455 | Study Design |  |  |  |  |  |
|  | 1500-1600 | WIPs, good supervisor, scholarly projects, struggles with project |  |  |  |  |  |
| Sept 25 Fri |  |  |  |  |  |  |  |
|  | 0805-0930 | Library- search strategies |  |  |  |  |  |
|  | 1000-1100 | How to use Zotero- you must have **Firefox** |  |  |  |  |  |
|  | 1100-1200 | Study Design |  |  |  |  |  |
|  | 1300-  1530 | Discuss – Critical Appraisal, WIPS, Good Supervisor, Project Struggles |  |  |  |  |  |
| Sept 28 Mon |  |  |  |  |  |  |  |
|  | 0800-0810 | Review literature search in the group |  |  |  |  |  |
|  | 0810-0955 | Confounders |  |  |  |  |  |
|  | 1010-1145 | Introductory stats didactic session |  |  |  |  |  |
|  | 1145-1150 | Search and Learn |  |  |  |  |  |
|  | 1315-1445 | CATS |  |  |  |  |  |
|  | 1445-1600 | First journal article prep (Groups) |  |  |  |  |  |
| Sept 29  Tues |  |  |  |  |  |  |  |
|  | 0805-0930 | Data Entry & Excel |  |  |  |  |  |
|  | 1000-1100 | Why do analyses and how to do them? |  |  |  |  |  |
|  | 1100-1200 | Critique j article in groups-hr #2 |  |  |  |  |  |
|  | 1300-1530 | Teaching tips and practice |  |  |  |  |  |
|  | Own time | IHI open school QI course complete  Part 3 of QI 101, full courses for QI102 + 103 (both) |  |  |  |  |  |
| Sept 30 Wed |  |  |  |  |  |  |  |
|  | 0805-1015 | *2 h time for IHI or TCPS2 or journal article or work on CAT* |  |  |  |  |  |
|  | 1030-1200 | Statistical -nuts and bolts |  |  |  |  |  |
| Oct 1 Thurs |  |  |  |  |  |  |  |
|  | 915-1150 | 3 journal articles – max 30-35 min and 10 min discussion and feedback |  |  |  |  |  |
|  | 1315-1420 | QI – Introduction |  |  |  |  |  |
|  | 1420-1640 | Feedback |  |  |  |  |  |
| Oct 2 Fri |  |  |  |  |  |  |  |
|  | 0805-1030 | Stats tutorial #2 |  |  |  |  |  |
|  | 1030-1115 | 4^th^ group journal article |  |  |  |  |  |
|  | 1230-1300 | *Resident to pick a topic with the ASK Chair or Karen at this time- (i.e. no advance planning)*  *- 30 min to search topic -prepare overview of literature as if speaking to a parent* |  |  |  |  |  |
|  | 1300-1530 | Search and learn |  |  |  |  |  |
| Oct 5  Mon |  |  |  |  |  |  |  |
|  | 0800-0930 | QI |  |  |  |  |  |
|  | 1000-1200 | Sample Size: Lecture and lab |  |  |  |  |  |
|  | 1315-1600 | Diagnostic testing |  |  |  |  |  |
| Oct 6  Tues |  |  |  |  |  |  |  |
|  | 0805-0830 | MCHP |  |  |  |  |  |
|  | 0900-1100 | PDSA cycles presentations  5 min per presentation and 5 min feeback. (1.5 h) |  |  |  |  |  |
|  | 1100-1200 | Systematic reviews- Kelly systematic review and Prisma guidelines |  |  |  |  |  |
|  | 1300-1545 | Qualitive research |  |  |  |  |  |
| Oct 7 Wed |  |  |  |  |  |  |  |
|  | 0805-0900 | Stats Hep |  |  |  |  |  |
|  | 0915-1015 | Ethics |  |  |  |  |  |
|  | 1005-1130 | Start to work on CAT, 2^nd^ journal article |  |  |  |  |  |
|  | After AHD – 1540 | 4 or 5 PGY4s to discuss how their scholarly projects went |  |  |  |  |  |
| Oct 8 Thurs |  |  |  |  |  |  |  |
|  | 0930-1100 | QI in practice |  |  |  |  |  |
|  | 1100-1200 | Qualitive article critique |  |  |  |  |  |
|  | 1320-1600 | 4 CATS |  |  |  |  |  |
| Oct 9 Fri |  |  |  |  |  |  |  |
|  | 0805-1000 | 4 CATS |  |  |  |  |  |
|  | 1000-1145 | Time management |  |  |  |  |  |
|  | 1330-1430 | Mixed Methods |  |  |  |  |  |
|  | 1430-1530 | Time to critique article 2 |  |  |  |  |  |

| Oct 13 Tues |  |  |
| --- | --- | --- |
|  | 0800-0900 | 2 CATS |
|  | 0900-1100 | Statistical models (regression) lecture., Ch 13-14 in Primer. |
|  | 1100-1200 | CHRIM- Clinical Research Unit (CRU) |
|  | 1300-1330 | Introductory talk about Lab Medicine |
|  | 1330-1500 | CV Session |
|  | 1515-1600 | Additional discussion about additional academic career training |
| Oct 14  Wed |  |  |
|  | 0805-0835 | ABDI |
|  | 0900-1100 | Present 2 journal articles |
|  | Post AHD | Meet with more senior residents who have worked on their projects |
| Oct 15  Thurs |  |  |
|  | 0915-1145 | Statistical models (regression) lab #3 |
|  | 1315-1400 | Poster session |
|  | 1400-1600 | 2 journal articles |
| Oct 16 Fri |  |  |
|  | 0800-1000 | Present 2 article |
|  | 1000-1100 | Ethics – Consent Issues |
|  | 1100-1200 | Create abstract |
|  | 1300-1500 | Presentation design, poster design, RSAC, Grants |
| Oct 19  Mon |  |  |
|  | 0800-0900 | Knowledge translation (KT) |
|  | 0915-1030 | How to prepare a manuscript |
|  | 1100-1150 | Help with regression homework |
|  | 1300-1500 | 2 articles presented by individual residents |
| Oct 20  Tues |  |  |
|  | 0805-1000 | 2 journal articles presenter |
|  | 1030-1200 | Team competition prep time |
|  | 1300-1500 | Competition #1 |
|  | 1500- | Review abstract with partners |
| Oct 21  Wed |  |  |
|  | 0800-1000 | Review of regression homework and more examples 5-10 |
|  | 1000-1100 | Present 1 article |

| Additions or other comments: |
| --- |
